# Optimizing Sepsis Treatment via Latent Protocol Inference

**DOI:** 10.64898/2026.11.23.26368780

**Authors:** Chenhui Wang, Vincent Liu, Edouardo Honig, Deqian Kong, Ying Nian Wu

**Affiliations:** Department of Statistics and Data Science, UCLA, Los Angeles, USA

## Abstract

Sepsis is a life-threatening medical emergency characterized by high mortality and variable treatment practices. Optimizing treatment from retrospective data is challenging due to sparse, delayed outcomes and complex physiological dynamics. We introduce the Generative Latent Protocol Model (GLPM), a probabilistic framework that structures clinical decision-making through a latent *protocol* variable anchored on the patient’s admission state. This latent variable acts as an information bottleneck, decoupling the generation of treatment trajectories from the prediction of survival outcomes. Unlike methods that rely on value estimation or noisy return-to-go conditioning, our model is trained via Maximum Likelihood Estimation to capture the joint distribution of treatment trajectories and outcomes. At inference time, we treat treatment optimization as a probabilistic inverse problem: we sample latent protocols from a posterior distribution conditioned on a target survival probability. On the ICU-Sepsis benchmark derived from MIMIC-III, our best variant matches or exceeds eight offline baselines on all five datasets, with gains that are statistically significant against every baseline on the most heterogeneous dataset, and it offers interpretability through patient-specific counterfactual analysis.

## 1 Introduction

Sepsis, defined as “life-threatening organ dysfunction caused by a dysregulated host response to infection” (Singer et al. 2016), contributes to at least 1.7 million adult hospitalizations and 350,000 deaths annually in the US (Dantes et al. 2023). Despite its prevalence, treatment guidelines for high-stakes interventions—such as intravenous fluids and vasopressors—remain inconclusive, leading to highly variable practice patterns and suboptimal outcomes (Byrne and Van Haren 2017). This lack of conclusive guidance indicates a critical need for unified, data-driven tools (Komorowski et al. 2018).

The sequential nature of sepsis care naturally fits the Markov decision process (MDP) framework. As illustrated in Figure 1, the whole treatment process of a patient can be viewed as a trajectory, a state–action sequence where the outcome (survival) is observed only at the end. Although reinforcement learning (RL) has been widely applied to this domain (Raghu et al. 2017; Komorowski et al. 2018; Peng et al. 2018; Choi et al. 2024), standard online methods face significant challenges in retrospective settings, including estimation bias and the difficulty of reliable off-policy evaluation (Jeter et al. 2019).

**Figure 1:**
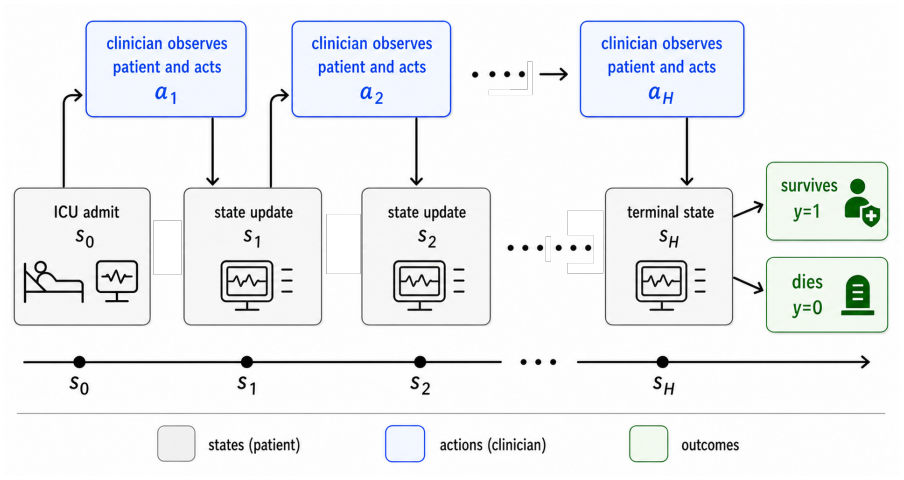
Sepsis treatment as sequential decision making. At each step the clinician observes the patient status and acts. The episode ends in survival (*y* = 1) or death (*y* = 0).

Recent work has shifted toward offline, trajectory-centric learning, framing healthcare as sequence modeling (Liventsev and Fritz 2024). Algorithms such as Diffuser (Janner et al. 2022) and Decision Transformer (DT) (Chen et al. 2021) have shown promise by modeling trajectories directly. However, in the context of sepsis treatment, these methods struggle with two key limitations: (1) *sparse, delayed rewards*, making credit assignment difficult for return-conditioned models in binary-outcome settings; and (2) *lack of temporal abstraction*, as flat policies often fail to capture the hierarchical, high-level structure of clinical care.

In this work, we start from a generative modeling perspective. We hypothesize that effective clinical strategies lie on a low-dimensional manifold embedded within the highdimensional space of patient trajectories. We capture this structure using a latent variable *z*, which we view as the underlying clinical *treatment protocol*: a learned representation, rather than a named clinical guideline. Instead of learning a fixed policy mapping states to actions, we learn a joint distribution over latents, trajectories, and outcomes. Our model follows the Latent Plan Transformer (LPT) (Kong et al. 2024b) in linking trajectories and outcomes through a shared latent variable. Generative Actor Critic (Qin et al. 2025) conditions its latent prior on the initial state and uses separate latent components for the trajectory and return. We condition on the patient’s admission state *s*_0_ and use a single protocol to generate treatment actions and predict a binary survival outcome.

This probabilistic formulation enables a robust mechanism for personalized treatment planning that operates on two distinct timescales. First, we perform global learning to capture general medical knowledge. By training on the entire population via Maximum Likelihood Estimation (MLE), the model learns population-level regularities of treatment and outcome. Second, we enable test-time adaptation (or patient-specific planning) for individual cases. Rather than applying a fixed policy, we treat the latent protocol *z* as a tunable parameter. For a new patient, we perform an inference step to identify the specific protocol *z* from the patient’s admission state to maximize the probability of survival. This allows us to “plan” via inference, effectively tailoring the strategy to the specific patient without requiring gradient updates to the model weights.

### Contributions

Our main contributions are:

#### 1 Generative Latent Protocol Mode

We introduce an *s*_0_-anchored hierarchical sequence model that captures treatment strategies via a latent bottleneck, trained by an Expectation-Maximization (EM) procedure on trajectory–outcome pairs.

#### 2 Goal-conditioned posterior inference

We formulate treatment recommendation as a probabilistic inference problem, and demonstrate that sampling from the survival-conditioned posterior allows for precise control over treatment aggressiveness.

#### 3 State-of-the-Art performance

On the ICU-Sepsis benchmark derived from MIMIC-III, our method consistently outperforms offline baselines across datasets of varying quality.

#### 4 Interpretability

We demonstrate that the learned latent space captures clinically meaningful treatment phenotypes (e.g., “wet” versus “dry” fluid strategies) and supports counterfactual analysis to identify missed interventions in non-survivor trajectories.

## 2 Related Work

### Decision making via sequence modeling

Conventional RL approaches optimize a policy *π*(*a*_t_|*s*_t*−*1_) to maximize expected return. However, sepsis treatment data is characterized by sparse, delayed rewards (outcome observed only at *t* = *H*) and complex physiological dynamics, making credit assignment difficult for standard value-based methods. Recent approaches have reframed decision-making as a sequence modeling problem. For instance, Decision Transformer (DT) employs an autoregressive Transformer to condition actions on desired returns and past trajectories (Chen et al. 2021), and the concurrent Trajectory Transformer plans by beam search (Janner, Li, and Levine 2021). Subsequent variants of DT include multi-task learning (Lee et al. 2022), online adaptation (Zheng, Zhang, and Grover 2022), and Q-function augmentation (Yamagata, Khalil, and Santos-Rodriguez 2023). Parallel to this, generative modeling has been applied to planning, most notably through diffusion-based trajectory generation (Ho, Jain, and Abbeel 2020; Janner et al. 2022); more recent planners make cross-trajectory stitching an explicit design objective (Luo et al. 2025).

While prior works like DT condition on return-to-go (RTG), we argue that RTG is a noisy signal in binary-outcome settings. Moreover, return-conditioned models often struggle in stochastic environments, where a high return may come from luck rather than from the actions taken (Yang et al. 2023; Paster, McIlraith, and Ba 2022). Latent variable models instead condition policies on latent variables, often learned from sub-trajectories (Lynch et al. 2020; Ajay et al. 2021). Our work follows this trajectory-centric perspective but differs in two key ways: it introduces latent representations of treatment protocols and a principled MLE-based training strategy, using an EM-style training procedure to infer episode-consistent protocol latent variables that provide temporal abstraction.

### Constraint-based and risk-sensitive RL

Our approach of conditioning on target survival rates during inference draws from constrained and risk-sensitive RL. Constrained MDPs (Achiam et al. 2017) separate optimization objectives from safety constraints, while distributional RL (Bellemare, Dabney, and Munos 2017; Dabney et al. 2018) and risk-constrained methods (Tamar, Glassner, and Mannor 2015; Chow et al. 2018) enable policies tailored to specific percentiles of outcome distributions. These frameworks motivated our exploration of different target survival rates to balance safety and treatment efficacy. As was evident, our findings aligned with theoretical insights, demonstrating that achievable high-probability constraints yield better outcomes than theoretical perfection in stochastic clinical environments.

### Latent variable sequence modelling

Latent Adaptive Planner updates a latent plan as new observations arrive during manipulation (Noh et al. 2025). In spatial planning, multi-horizon transition representations guide action selection toward a goal (Zhao et al. 2025; Kong et al. 2026a). In our setting, an episode-level latent represents the treatment protocol and is inferred before executing the policy. Related sequence models use latent initial states to generate continuoustime dynamics (Cheng et al. 2025) and latent thought vectors to guide language generation and revise reasoning traces (Kong et al. 2025, 2026b). A critical component of these frameworks is the mechanism for posterior inference and exploration. While variational methods like VariBAD provide computationally efficient, amortized inference (Zintgraf et al. 2021), posterior sampling offers a principled alternative for balancing exploration and exploitation (Osband, Russo, and Van Roy 2013). When exact posterior sampling is infeasible, Langevin dynamics offer a practical approximation with provable guarantees for both online exploration (Ishfaq et al. 2024) and offline RL (Nguyen-Tang et al. 2024). Building on this foundation, we investigate Langevin sampling and variational inference to balance uncertainty quantification with computational constraints.

## 3 Methods

### 3.1 Sepsis Treatment as Sequential Decision Making

We formulate sepsis treatment as a finite-horizon sequential decision-making problem. A patient trajectory is denoted by *τ* = (*s*_0_, *a*_1_, *s*_1_, *a*_2_, *s*_2_, … , *a*_H_ , *s*_H_), where *s*_t_ ∈ *S* represents the physiological state, *a*_t_ ∈ A denotes the clinical intervention, and *H* is the horizon. Each trajectory is associated with a binary terminal outcome *y* ∈ {0, 1}, indicating patient survival. Define the history *h*_t_ = (*s*_0:t*−*1_, *a*_1:t*−*1_), with the policy keeping only the most recent *K* steps. We learn from an offline dataset of trajectory–outcome pairs (*τ*_i_, *y*_i_) and infer a latent *protocol* from a new initial state and a desired outcome at test time.

### 3.2 Generative Latent Protocol Model

We propose a generative model that treats the clinical course and the patient outcome as conditionally independent given a latent *protocol z* ∈ ℝ^d^. This latent variable acts as an information bottleneck, capturing the high-level medical strategy governing the episode. A similar factorization links a generated object and its predicted properties through a shared latent in molecular design (Kong et al. 2023, 2024a). The joint distribution is defined as:

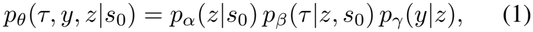

where *θ* = (*α, β, γ*) are learnable parameters. The generative process is structured as follows (see Figure 2):

**Figure 2:**
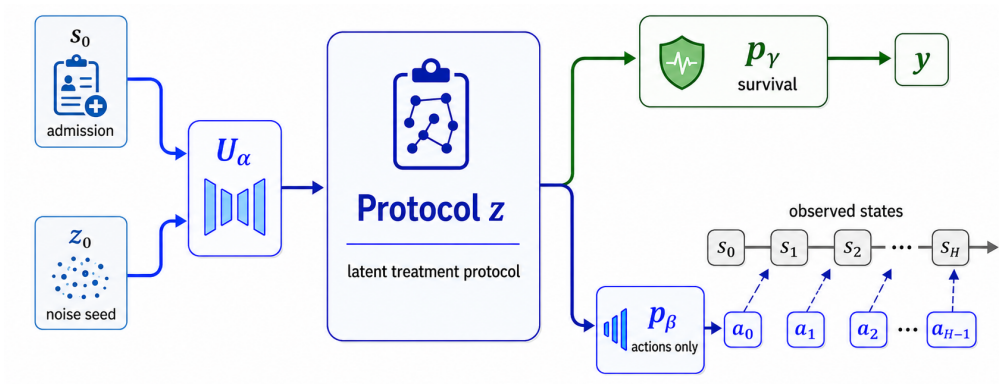
Overview of Generative Latent Protocol Model. A latent protocol *z* ∈ ℝ^d^ is obtained via neural transform *z* = *U*_α_(*z*_0_, *s*_0_) with *z*_0_ ∼ *N* (0, *I*_d_). Conditioned on *z*, the trajectory generator *p*_β_(*τ* |*z, s*_0_) and survival predictor *p*_γ_(*y*|*z*) model the trajectory *τ* and terminal outcome *y*, respectively.

The prior model *p*_α_(*z*|*s*_0_) is an implicit generator, *z* = *U*_α_(*z*_0_, *s*_0_) with *z*_0_ ∼ *p*_0_ = *N* (0, *I*_d_) (Kong et al. 2024a,b). Here, *U*_α_ is a non-linear network (a U-Net (Ronneberger, Fischer, and Brox 2015)), allowing the model to learn a complex manifold of valid treatment protocols from a simple base distribution. The trajectory generator *p*_β_(*τ* |*z, s*_0_) is an autoregressive model with a finite context window of *K* steps, 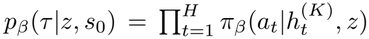, where 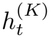 contains the *K* most recent states and actions of the history *h*_t_ (truncated at the episode start for *t < K*). We realize the generator using a causal Transformer in which *z* enters through cross-attention, similar to DT (Chen et al. 2021). The relationship between a protocol and patient survival is modeled by a Bernoulli predictor *p*_γ_(*y*|*z*) = *f*_γ_(*z*)^y^[1 − *f*_γ_(*z*)]^1*−*y^ with logit *s*_γ_(*z*) and survival probability *f*_γ_(*z*) = sigmoid(*s*_γ_(*z*)), computed by a small multilayer perceptron (MLP).

### 3.3 Maximum likelihood learning

Given the offline dataset 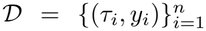 , we learn *θ* by maximizing the marginal log-likelihood 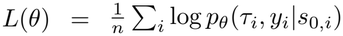. Direct optimization of *L*(*θ*) is intractable due to the integral over the latent space. However, using the Fisher identity (Section A), the gradient can be expressed as an expectation over the posterior *p*_θ_(*z*_0_|*τ, y, s*_0_):

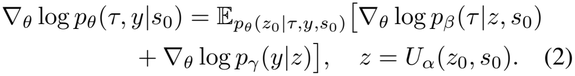

This formulation naturally suggests an iterative Expectation-Maximization (EM) learning scheme:

- **Inference (E-step):** For each trajectory–outcome pair (*τ, y*) with admission state *s*_0_, we infer the posterior distribution of the latent noise *p*_θ_(*z*_0_|*τ, y, s*_0_), and let *z* = *U*_α_(*z*_0_, *s*_0_). This step identifies the latent protocol *z* that best explains the observed clinical decisions and patient outcome.
- **Learning (M-step):** We update the parameters *θ* via stochastic gradient ascent to maximize the joint likeli-hood given the inferred protocols.

The core computational challenge lies in the E-step. The posterior distribution *p*_θ_(*z*_0_|*τ, y, s*_0_) is proportional to the product of the prior and the likelihoods:

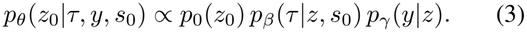

This distribution is highly non-Gaussian and multimodal. To facilitate sampling via Langevin dynamics, we characterize the posterior by its energy function *E*(*z*_0_), such that *p*_θ_(*z*_0_|*τ, y, s*_0_) ∝ exp{−*E*(*z*_0_)}:

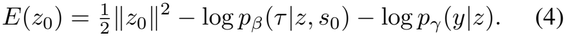

Exact sampling from this distribution is intractable. Therefore, we rely on approximate posterior sampling strategies to estimate the expectation in Equation (2).

### 3.4 Fast–slow adaptation mechanism

This EM-style framework separates fast inference of local latents from slow learning of shared weights, as in latent thought models (Kong et al. 2025). A related meta-learning approach adapts a latent-space energy-based prior to each new task while holding the generator fixed (Kong, Pang, and Wu 2021). We distinguish between two sets of parameters:

- **Global “slow” parameters (***θ*): The neural network weights (*α, β, γ*) are shared across the entire population. They capture population-level regularities of treatment and outcome, learned slowly over the entire dataset.
- **Local “fast” parameters (***z*): The latent protocol *z* acts as a patient-specific parameter vector. During inference (the E-step), *z* is optimized specifically for the current episode (*τ*_i_, *y*_i_).

This separation enables single-instance adaptation. By optimizing *z* via posterior sampling, the model effectively “finetunes” the policy’s behavior to the nuances of a specific patient without modifying the global network *θ*. This allows for personalized treatment planning at test time while keeping the robustness of a population-level model.

### 3.5 Posterior inference strategies

The gradient estimator in Equation (2) relies on drawing samples from the posterior distribution defined by the energy function *E*(*z*_0_) (Equation (4)). Short-run Langevin sampling has also been used to infer continuous response latents in dialogue models (Xu et al. 2023). We investigate two E-step algorithms to approximate this expectation, offering different trade-offs between mixing accuracy, computational cost, and implementation complexity.

#### BAOAB

Standard overdamped dynamics behave as a random walk, which can suffer from slow mixing in high-dimensional latent spaces. To sample the underdamped Langevin dynamics on *E*(*z*_0_) numerically, we adopt the BAOAB integrator (Leimkuhler and Matthews 2013), which splits the dynamics into Force (B), Drift (A), and Ornstein– Uhlenbeck (O) steps, applied symmetrically. Using step size *h*, friction *ζ*, momentum *v*, and noise sample *ϵ*:

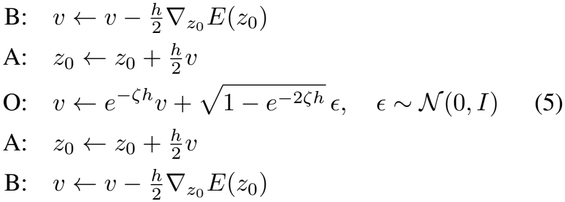

The symmetric composition gives second-order accuracy of the sampled distribution, allowing for larger step sizes *h* than standard solvers while maintaining stability. The local state is the chain position and momentum 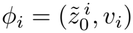.

### Stochastic variational inference (SVI)

As an alternative to MCMC sampling, we employ stochastic variational inference by approximating the true posterior *p*_θ_(*z*_0_|*τ, y, s*_0_) with a diagonal Gaussian *q*_ϕ_(*z*_0_) = *N* (*µ*, diag(*σ*^2^)), where the variational parameters *ϕ* = (*µ, σ*) are treated as local parameters specific to each trajectory. Instead of training a separate inference network (amortization), we directly optimize *µ* and *σ* for the current episode to maximize the evidence lower bound (ELBO):

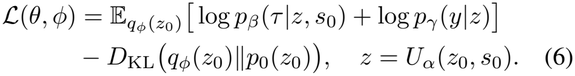

Gradients are estimated via the reparameterization trick (Kingma and Welling 2014), *z*_0_ = *µ* + *σ* ⊙ *ϵ* with *ϵ* ∼ N (0, *I*). We perform this local optimization using the Adam optimizer (Kingma and Ba 2015). This transforms the E-step into a distinct optimization problem: rather than sampling chains, we find the optimal Gaussian approximation of the protocol posterior for the specific patient.

### 3.6 Protocol-conditioned treatment

Once trained, the generative model functions as a goal-conditioned policy. We generate treatment trajectories conditioned on a prospective survival target *y*^⋆^ ∈ [0, 1] and the admission state:

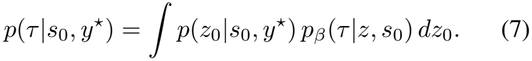

Inference proceeds in two stages: (1) planning, where we infer a latent protocol *z* that satisfies the survival target; and (2) execution, where we generate an action sequence conditioned on this protocol.

#### Stage 1: Latent protocol planning

We infer the latent protocol by sampling *z*_0_ from the target-conditioned posterior *p*(*z*_0_|*s*_0_, *y*^⋆^) ∝ *p*_0_(*z*_0_) *p*_γ_(*y*^⋆^|*z*). Unlike the training posterior, which is constrained by an observed trajectory, this posterior favors regions of the latent manifold where the predictor assigns high probability to the target outcome under the prior. While the model is trained on binary outcomes, clinical goals are often best expressed as a continuous target probability (e.g., *y*^⋆^ = 0.95). We therefore tilt the prior by the binary cross-entropy with a temperature *T*_y_ *>* 0, giving the planning energy

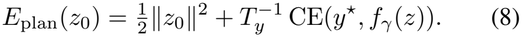

Minimizing the cross-entropy is equivalent to minimizing the KL divergence between a target Bernoulli distribution Q(*y*^⋆^) (the desired risk profile) and the predicted P(*f*_γ_(*z*)). This formulation allows us to continuously tune the aggressiveness of the treatment protocol by varying *y*^⋆^ within the unit interval. The gradient of the planning energy is

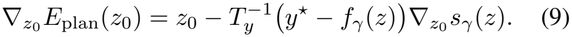

To sample from this posterior, we utilize the same strategies described in Section 3.5, substituting the training energy *E*(*z*_0_) with the planning energy *E*_plan_(*z*_0_). The resulting sample *z*^⋆^ represents an optimized treatment protocol.

#### Stage 2: Action execution

Given *z*^⋆^, execution is purely autoregressive. At each timestep *t*, the agent samples actions from the policy conditioned on the fixed protocol: 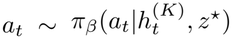. By keeping *z*^⋆^ fixed throughout the episode, GLPM ensures that individual decisions remain temporally coherent and aligned with the high-level survival strategy identified during the planning stage.

## 4 Experiments

### 4.1 Setup

To evaluate our proposed method, we utilize the ICU-Sepsis environment (Choudhary, Gupta, and Thomas 2024), a standardized tabular MDP for sepsis management constructed from the records of approximately 17,000 sepsis patients in MIMIC-III (Johnson et al. 2016). This environment poses a challenging sequential decision-making task with 716 discrete states, a sparse, binary survival outcome, and 25 discrete treatment combinations, representing discretized dosages of IV fluids and vasopressors (Table 6, Section C).

#### Offline dataset construction

Given the lack of standardized offline RL benchmarks for ICU-Sepsis, we constructed a suite of datasets following the D4RL methodology (Fu et al. 2020). We generated behavior policies of varying quality by training Soft Actor-Critic (SAC) agents online and capturing checkpoints at different performance thresholds. **Random**: 1M steps collected by a randomly initialized policy. **Medium-Replay**: the full replay buffer of a SAC agent trained until medium-level performance. **Medium**: 1M steps from a fixed, partially trained policy. **Expert**: 1M steps from a fully converged SAC policy (about 83% survival). **Real**: real-world clinical trajectories from MIMIC-III, mapped into the ICU-Sepsis MDP following Komorowski et al. (2018).

#### Baselines

We compare against representative offline baselines: Behavior Cloning (BC) (Atkeson and Schaal 1997), Conservative Q-Learning (CQL) (Kumar et al. 2020), DT (Chen et al. 2021), Q-Learning Decision Transformer (QDT) (Yamagata, Khalil, and Santos-Rodriguez 2023), Trajectory Transformer (TT) (Janner, Li, and Levine 2021), Diffuser (Janner et al. 2022), Decision Diffuser (DD) (Ajay et al. 2023), and LPT (Kong et al. 2024b). Baseline implementations were adapted from official code. All policies are evaluated in the ICU-Sepsis environment, and results are averaged over five random seeds. GLPM infers each latent protocol with target survival rate *y*^⋆^ = 0.95; Section D lists all settings.

### 4.2 Main results

Table 1 details the comparative performance in the offline setting. No baseline surpasses our best variant on any dataset: BAOAB achieves the highest survival rate on Medium-Replay, Medium, and Real and ties for best on Expert, while SVI ties for the best result on the low-quality Random data. Among the baselines, LPT is the strongest on every regime except Real, where QDT leads, and DD degrades on Expert (80.6%), consistent with the cluster collapse of classifier-free guidance at narrow return distributions (Section E).

**Table 1:** Performance on ICU-Sepsis (mean ± std survival, %). All policies are evaluated in ICU-Sepsis. Results are averaged over 5 seeds; the best results are in **bold** and the second best are <u>underlined</u>.

| Algorithm | Random | Med.-Replay | Medium | Expert | Real |
|---|---|---|---|---|---|
| BC | 79.80±3.66 | 80.00±4.34 | 84.00±2.68 | 88.00±2.17 | 79.40±3.44 |
| CQL | 76.60±3.72 | 80.60±4.22 | 79.20±3.97 | 77.60±2.42 | 78.40±3.55 |
| DT | 78.20±3.25 | 79.20±2.14 | 84.00±2.97 | 87.40±3.93 | 80.20±2.93 |
| QDT | 80.80±4.17 | 83.00±1.67 | 86.00±1.10 | 88.00±2.61 | 80.40±4.13 |
| TT | 79.20±6.70 | 81.80±4.10 | 83.30±2.70 | 83.90±3.60 | 79.70±4.20 |
| Diffuser | 77.00±5.60 | 81.20±1.90 | 82.60±1.90 | 86.00±1.20 | 79.80±3.00 |
| DD | 78.60±3.00 | 80.40±3.50 | 80.00±2.40 | 80.60±3.00 | 78.20±1.30 |
| LPT | **81.60±3.01** | 87.00±3.36 | 87.60±2.42 | **91.00±2.41** | 80.20±1.17 |
| GLPM (BAOAB) | 80.50±2.77 | **89.00±3.13** | **89.00±4.00** | **91.00±2.17** | **83.20±2.68** |
| GLPM (SVI) | **81.60±6.02** | 85.00±2.68 | 87.00±1.79 | 88.00±2.10 | 82.00±4.47 |

On Medium-Replay and Medium, BAOAB achieves 89.0% on both, ahead of the strongest baseline by 2.0 and 1.4 points respectively and well clear of the return-conditioned and generative-planning baselines (QDT 83.0%/86.0%; TT 81.8%/83.3%). It also exceeds the online behavior policies used to collect the data. The average margin over the baselines is largest on Medium-Replay, the most hetero-geneous dataset; we attribute this to a trajectory-stitching effect discussed in Section 5. On Expert, BAOAB reaches 91.0%, matching the best baseline and clearing the 88.0% of BC and QDT (best diffusion planner: Diffuser, 86.0%). On Real, which arguably presents the hardest challenge due to noise and confounding, BAOAB achieves 83.2%, against the ≈ 80% reached by every baseline (best: QDT, 80.4%). Overall, the BAOAB integrator yields the highest-performing policies as dataset quality and heterogeneity increase, with SVI competitive on the low-quality Random regime.

#### Statistical significance

Because several baseline standard deviations overlap, we quantify the BAOAB gains with a paired bootstrap (Table 2). On Medium-Replay, the trajectory-stitching regime, BAOAB’s gain is significant against all baselines (*p <* 0.05). Across the full set, the breadth of significance tracks each baseline’s strength: BAOAB significantly exceeds CQL on four of five datasets, BC and DD on three, TT and Diffuser on two, and DT, QDT, and LPT on Medium-Replay alone. Against QDT, the gains on Medium, Expert, and Real are consistent in direction (+2.8 to +3.0 points) but do not reach significance at five seeds. LPT is the hardest to separate from: BAOAB’s only significant margin over it is the +2.0 points on Medium-Replay (*p* = 0.048), and the two tie on Expert. Because LPT already adopts a return-free latent abstraction in place of return-to-go, we attribute these residual differences to our *s*_0_-anchored prior and the better-mixing BAOAB chain, which surface in the heterogeneous Medium-Replay regime where trajectory stitching is most valuable.

**Table 2:** Paired-bootstrap comparisons of BAOAB with each baseline: BAOAB gain in percentage points /two-sided *p*-value, using 10,000 resamples. **Bold** marks *p <* 0.05.

| Algorithm | Random | Med.-Replay | Medium | Expert | Real |
|---|---|---|---|---|---|
| BC | +0.7/.38 | +9.0/<**.001** | +5.0/**.048** | +3.0/**.026** | +3.8/.083 |
| CQL | +3.9/.08 | +8.4/**.005** | +9.8/**.002** | +13.4/<**.001** | +4.8/**.049** |
| DT | +2.3/.18 | +9.8/<**.001** | +5.0/.054 | +3.6/.055 | +3.0/.120 |
| QDT | −0.3/.56 | +6.0/<**.001** | +3.0/.098 | +3.0/.094 | +2.8/.183 |
| TT | +1.3/.12 | +7.2/**.003** | +5.7/.062 | +7.1/**.013** | +3.5/.073 |
| Diffuser | +3.5/.092 | +7.8/<**.001** | +6.4/**.015** | +5.0/.057 | +3.4/.066 |
| DD | +1.9/.18 | +8.6/<**.001** | +9.0/**.005** | +10.4/<**.001** | +5.0/.051 |
| LPT | −1.1/.48 | +2.0/**.048** | +1.4/.082 | +0.0/.62 | +3.0/.157 |

#### Comparison with online RL

As a performance reference, agents trained online directly on the environment, a Stochastic Latent Actor–Critic (Lee et al. 2020) and PPO (Schulman et al. 2017), reach final survival of 81% and 86%. Our offline BAOAB model (mean 86.5% over the five datasets, Expert 91.0%) is competitive with these online agents despite training without any environment interaction. Inference is also cheap relative to the clinical timescale: a full BAOAB episode takes 1.18 s on one GPU against a 4-hour decision interval (Section I).

### 4.3 Prospective target conditioning

A central feature of our framework is *prospective target conditioning*: the ability to specify desired survival rates *y*^⋆^ at inference time. This raises a practical question: which target yields the most effective strategy? We conducted an ablation study to evaluate the sensitivity of the model to different inference targets *y*^⋆^ ∈ {0.85, 0.90, 0.95, 1.00} as shown in Table 3. Across datasets, *y*^⋆^ = 0.95 consistently yields the best performance, while pushing the target to theoretical perfection (*y*^⋆^ = 1.00) degrades it. This suggests that *y*^⋆^ = 0.95 finds a safer, high-likelihood *protocol* that balances ambition with robustness, whereas *y*^⋆^ = 1.00 leaves the search dominated by the prior.

**Table 3:** Ablation study of performance across target survival rates (mean ± std, %). Results are averaged over 5 seeds; higher is better. **Bold** indicates the best result per dataset.

| Dataset | Target *y*<sup>⋆</sup> |  |  |  |
|---|---|---|---|---|
|  | 0.85 | 0.90 | 0.95 | 1.00 |
| Random | 80.00±5.10 | 77.80±3.06 | **80.50±2.77** | 79.60±4.03 |
| Med.-Replay | 86.20±2.32 | 87.40±3.44 | **89.00±3.13** | 84.80±2.32 |
| Medium | 82.30±1.83 | 84.00±2.97 | **89.00±4.00** | 85.80±3.49 |
| Expert | 86.40±3.98 | 85.40±4.59 | **91.00±2.17** | 88.00±3.16 |
| Real | 77.20±2.32 | 77.40±2.73 | **83.20±2.68** | 76.60±2.87 |
| Interv. level | Conserv. | Balanced | Balanced | Aggress. |

#### Optimization landscape

To explain why the extreme target underperforms, we visualize the gradient field 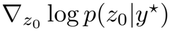 (Figure 3). For *y*^⋆^ = 1.00, the field becomes nearly radial and is dominated by contraction toward a single attractor: the target-conditioned posterior is largely pulled back toward prior-preferred regions. For *y*^⋆^ = 0.95, the field is no longer radial: it navigates *z*_0_ toward high-survival regions while staying anchored to the prior support. Equation (9) explains this difference. Its Gaussian term *z*_0_ pulls the code toward the origin, and its outcome term scales with *y*^⋆^ − *f*_γ_(*z*). Where the predictor saturates near one, the outcome term vanishes at *y*^⋆^ = 1.00 and leaves only the Gaussian pull. At *y*^⋆^ = 0.95, the outcome term changes sign wherever *f*_γ_ exceeds the target, so it keeps shaping the search. This suggests that GLPM performs best when searching for the best *realistic* protocol rather than an idealized outlier.

**Figure 3:**
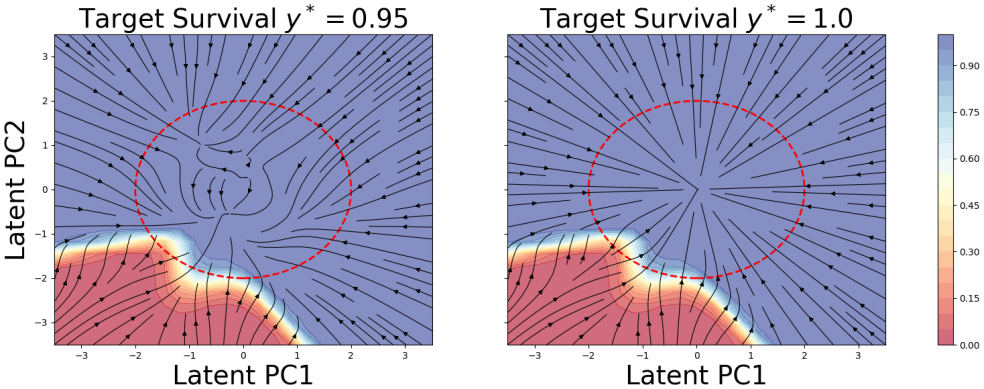
Gradient field 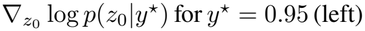 and *y*^⋆^ = 1.00 (right), projected onto the first two principal components of *z*_0_. Shading gives the predicted survival prob-ability. The dashed circle marks two standard deviations of the Gaussian prior.

### 4.4 E-step sensitivity and convergence

A natural concern is whether so few Langevin steps in the E-step yield a converged EM objective and a stable posterior. We vary the training-time Langevin steps *N* ∈ {1, 5, 10} on Medium-Replay. Over the ten EM iterations, the total negative log-likelihood falls steadily and flattens, and the posterior displacement Δ*z* stays flat, so each E-step moves the stored chain state by a nearly constant amount (Figure 7, Section F). Even one Langevin step per E-step thus gives a converging EM objective and a stable posterior. During testing, we keep the inference budget fixed at *N* = 25 (Table 4). Survival is highest at *N* = 5, while *N* = 1 under-runs the posterior and *N* = 10 brings no further gain. Δ*z* follows the same pattern: it grows from 12.6 to 15.8 between *N* = 1 and *N* = 5 steps and saturates at 16.0 with *N* = 10. Therefore *N* = 5 is a sufficient E-step depth.

**Table 4:** E-step depth ablation on Med.-Replay (mean ± std survival, %). Δ*z* is the per-iteration posterior displacement.

| Training steps | Survival | Displacement Δ*z* |
|---|---|---|
| 1 | 83.30±5.00 | 12.6 |
| **5** | **89.00±3.13** | 15.8 |
| 10 | 85.30±4.10 | 16.0 |

We further test eight BAOAB chains started from independent prior draws and find that they need at most 13 steps of burn-in, within the 25-step inference budget, and agree with Gelman–Rubin 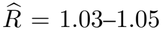 (Gelman and Rubin 1992), below the standard 1.1 threshold.

### 4.5 Context-length sensitivity

We set the policy context length of GLPM to *K* = 10, which is the mean trajectory length in the Medium-Replay dataset. A natural question is to ask how GLPM will perform if we change the context length. We vary *K* ∈ {5, 10, 20} and test BAOAB on Medium-Replay and keep every other hyperparameter at its default. Since DT also conditions on a window of the last *K* steps, we also include DT as a comparison. As the 90th percentile of the trajectory length is 21, *K* = 20 already covers whole trajectories in most cases.^1^ As shown in Table 5, BAOAB scores higher than DT at every *K*, by 7.0 to 9.8 points. Its survival is highest at *K* = 10; *K* = 5 lowers it to 85.3%, and *K* = 20 gives 87.2% with a wider spread across seeds. DT rises slowly with *K*, from 78.3% to 80.2%, but is always lower than BAOAB. The advantage of BAOAB therefore does not depend on the context length, and *K* = 10 is the best of the three settings for BAOAB.

**Table 5:**
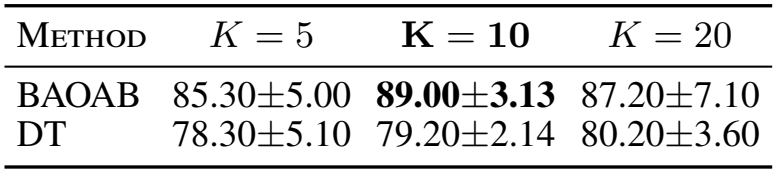
Context length *K* ablation on Med.-Replay for BAOAB and DT (mean ± std survival, %). **Bold** represents the best result.

| Method | *K* = 5 | **K = 10** | *K* = 20 |
|---|---|---|---|
| BAOAB | 85.30±5.00 | **89.00±3.13** | 87.20±7.10 |
| DT | 78.30±5.10 | 79.20±2.14 | 80.20±3.60 |

### 4.6 Qualitative analysis and interpretability

We examined the decision-making rationale of the generative model across three levels: latent space geometry, patientspecific counterfactuals, and population-level statistics.

#### Latent space geometry and controllability

To verify whether the latent space captures coherent clinical strategies, we linearly interpolate between two learned protocols with opposite fluid use: a “dry” protocol *z*_dry_ (low fluid doses) and a “wet” protocol *z*_wet_ (high fluid doses), *z*_λ_ = (1 − *λ*)*z*_dry_ + *λz*_wet_ for 0 ≤ *λ* ≤ 1, and compare the recommended doses over 15 time steps (Figure 4). The wet protocol administers fluids at *t* = 3, one step before the dry protocol, and the two agree at every other step. For vasopressors, at six of the fifteen steps the wet protocol suggests lower doses than the dry protocol, and it never recommends higher ones. Intermediate values of *λ* switch between the two protocol doses step by step instead of taking values in between. Moving from *z*_dry_ to *z*_wet_ therefore trades vasopressor support for earlier fluids: a single direction in the latent space couples the timing of fluid resuscitation with the level of vasopressor support.

**Figure 4:**
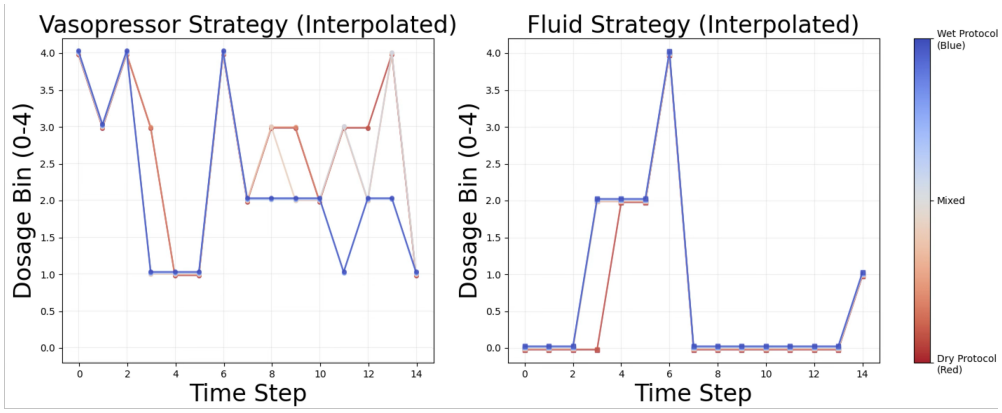
Recommended vasopressor (left) and fluid (right) levels along *z*_λ_, from the dry protocol (red, *λ* = 0) to the wet protocol (blue, *λ* = 1), over 15 time steps.

#### Preventing premature weaning

We compare GLPM’s recommendations with a clinician’s by replaying the record of Patient 27, a non-survivor from the test set with 33 four-hour steps. We infer *z*^⋆^ with the planning procedure of Section 3.6 at *y*^⋆^ = 0.95. GLPM diverges from the clinician at 7 of the 33 steps (21.2%; Figure 5), revealing two key differences: (1) *early fluid intervention*: GLPM starts fluids at *t* = 1, one step before the clinician’s large bolus at *t* = 2, and gives no fluid at *t* = 2; and (2) *de-escalation prevention*: after the clinician lowers the vasopressor dose from 4 to 1 at *t* = 10 and from 4 to 2 at *t* = 20, GLPM lowers it less and keeps it above the clinician’s dose for two steps each time, and when the clinician lowers it again from 2 to 1 at *t* = 21, GLPM keeps it at 3. Across the seven divergent steps, the net vasopressor change is +2 levels and the net fluid change is −2 levels. The model effectively corrects the reactive “rollercoaster” pattern of clinician care, favoring proactive stability.

**Figure 5:**
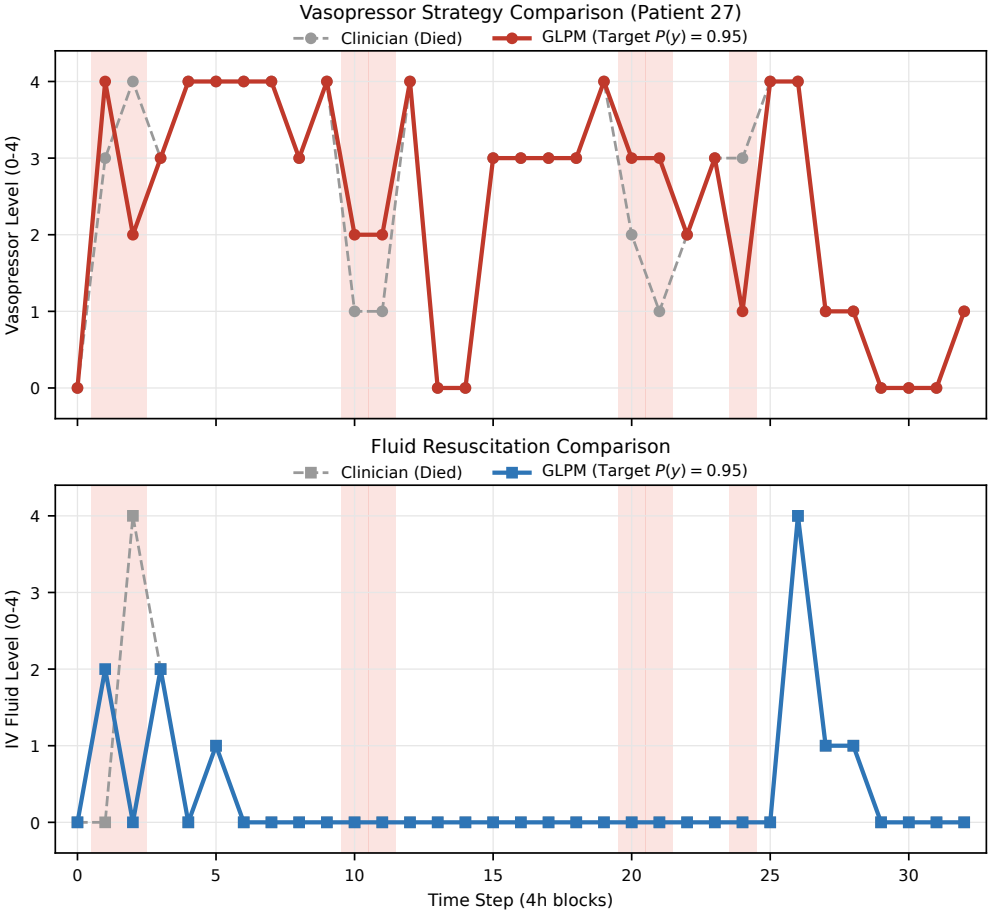
Counterfactual analysis of Patient 27 (non-survivor). *(Top)* Vasopressor strategy: GLPM (solid) maintains elevated support where the clinician (dashed) de-escalated prematurely. *(Bottom)* Fluid resuscitation: GLPM initiates earlier (*t* = 1), smoothing the intervention profile. Shading marks the seven divergent steps.

#### Generalization across non-survivors

Repeating the analysis for 50 further non-survivors with at least 20 steps (Section H), the divergence rate is 24.2% ± 9.8%: GLPM disagrees with the clinician at about one fourth of the steps. GLPM recommends higher vasopressor support, with a net change of +1.3 ± 4.5 levels, while the net fluid change is +0.1 ± 2.9 levels and is zero for most patients. De-escalation prevention occurs in 41 of the 50 patients, 1.8 ± 1.5 times per patient. The population behavior thus repeats the pattern of Patient 27: GLPM keeps vasopressor support higher and smoother than the clinician without giving more fluid.

#### Action distributions

We compare the intervention distributions of GLPM and DT, both trained on Real, with actual clinician behavior (Figure 6). A consistent pattern emerges: while clinicians favor lower vasopressor doses and spread fluid dosages across all five levels, both learned policies shift toward substantially higher vasopressor doses and give almost no fluid. Policies trained on Expert show the same shift (Section K). This identifies an alternative, more aggressive hemodynamic management strategy that correlates with higher survival in this environment.

**Figure 6:**
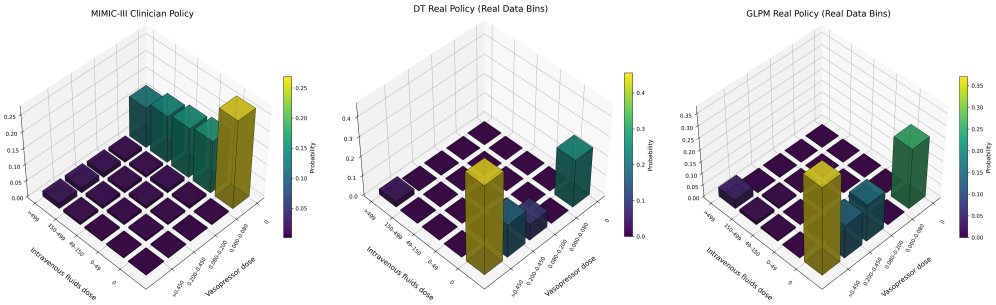
Action distributions under the dose bins of Table 6, aggregated over all time steps: clinicians in MIMIC-III (left), and DT (middle) and GLPM (right) trained on Real.

**Figure 7:**
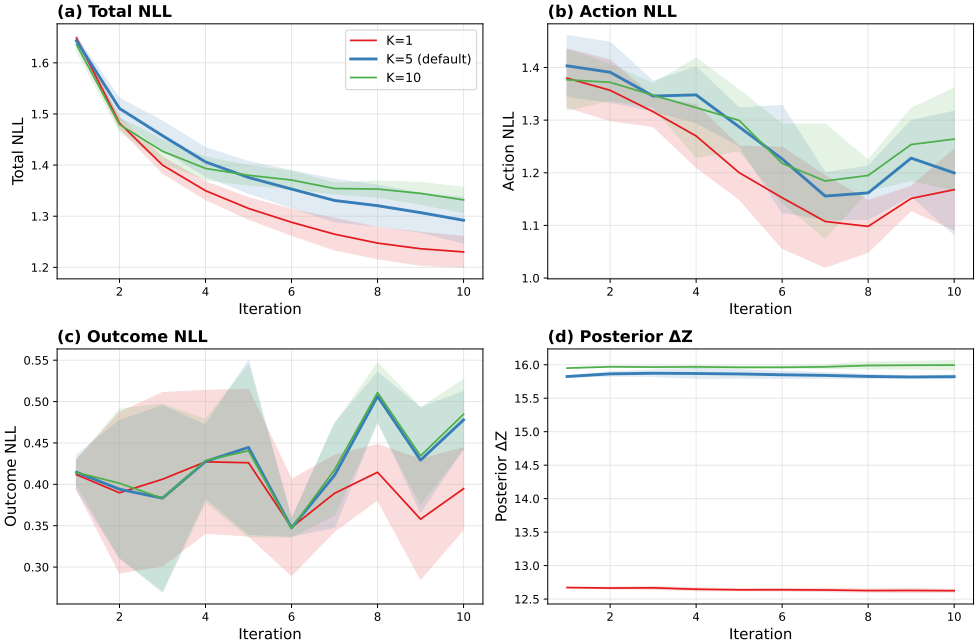
EM convergence on Medium-Replay for training E-step depths *N* ∈ {1, 5, 10} (labeled *K* in the legend). (a) Total, (b) action, and (c) outcome negative log-likelihood; (d) per-iteration posterior displacement Δ*z*.

## 5 Discussion

### Mechanism and advantages

Our method’s performance stems from two design choices: learning from full trajectory– outcome pairs via MLE and utilizing a latent variable *z* for temporal abstraction. Unlike behavior cloning, which suffers from compounding errors over long horizons (Ross, Gordon, and Bagnell 2011), our generative approach improves stability by guiding action generation via the protocol. Unlike CQL, DT variants, and diffusion-based planners, which can struggle with sparse rewards or noisy return-to-go targets (Paster, McIlraith, and Ba 2022; Brandfonbrener et al. 2022), our method avoids value estimation biases by directly optimizing the *s*_0_-anchored joint distribution. LPT shares our return-free latent-abstraction view; our gains over it stem from the *s*_0_-anchored prior and the better-mixing BAOAB chain.

### Why BAOAB wins on Medium-Replay

Medium-Replay yields our largest and statistically strongest margin, and it is also the most heterogeneous dataset: the replay buffer of a partially trained SAC agent mixes low-survival exploratory trajectories with the later high-survival trajectories that followed, so a method that composes useful fragments across trajectories of differing quality is well positioned. Brandfonbrener et al. (2022) show that return-conditioned supervised learning (DT and its variants) cannot stitch trajectories in general, even with unlimited data, because conditioning ties each generated trajectory to the empirical return of *individual* trajectories rather than to their optimal composition. Luo et al. (2025) instead make stitching the central design objective of a diffusion planner. LPT demonstrated trajectory stitching using a latent plan inferred from a target return (Kong et al. 2024b). Our Medium-Replay results are consistent with this account, though the benchmark comparison does not isolate stitching as the source of the gain.

### Clinician versus AI dosing

Qualitatively, our learned policies tended to recommend higher vasopressor doses and lower IV fluid volumes than clinicians (Figure 6). While consistent with prior observations of the AI Clinician (Komorowski et al. 2018), these tendencies reflect the dynamics of the simulation benchmark and require careful validation before real-world interpretation.

### Limitations and future directions

We acknowledge that ICU-Sepsis is a simplified tabular MDP with discrete states and a limited action space, preventing it from capturing the full physiological complexity of sepsis care or intermediate outcomes like organ recovery. Importance-sampling off-policy estimates on held-out Real trajectories are uninformative for policies that depart from clinician behavior (Section J), so our evaluation rests on the simulator. Future work will extend latent protocol inference to real-world cohorts with irregular sampling and confounding, such as MIMIC-IV (Johnson et al. 2023; Huang, Yang, and Rahmani 2025), and integrate these protocols into clinical decision support workflows (Sendak et al. 2020).

## 6 Conclusion

In summary, we introduced a latent protocol inference frame-work that provides temporal abstraction and enables goal-conditioned planning without reliance on step-wise rewards. Representing treatment strategies as latent vectors anchored on the admission state matches or exceeds strong offline baselines on every ICU-Sepsis dataset, offering a promising avenue for refining clinical decision-making through generative modeling.

## Data Availability

The ICU-Sepsis simulation environment used in this study is publicly available at https://github.com/icu-sepsis/icu-sepsis. Four offline datasets (Random, Medium-Replay, Medium, and Expert) were generated using behavior policies within this environment. The Real dataset consists of clinical trajectories derived from MIMIC-III. The original MIMIC-III database is available at https://physionet.org/content/mimiciii/1.4/ to credentialed researchers, subject to the applicable data use agreement.

https://physionet.org/content/mimiciii/1.4/

## Impact Statement

In the near term, the findings in this study should not be used to inform medical practice. Latent protocol inference is a methodological contribution that could inform future research on decision support for sepsis care, but any clinical application would require extensive prospective evaluation using real-time data and clinical trials.

## A Learning Gradient

Given training examples 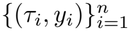 , the log-likelihood 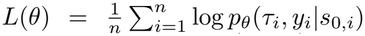. Dropping the index *i*, with *z* = *U*_α_(*z*_0_, *s*_0_) and *p*_θ_(*τ, y, z*_0_|*s*_0_) = *p*_0_(*z*_0_) *p*_β_(*τ* |*z, s*_0_) *p*_γ_(*y*|*z*),

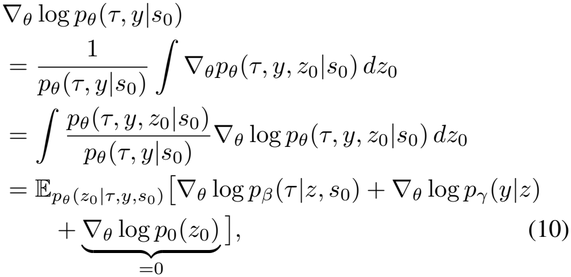

where the first line requires that differentiation can pass through the integral and the second uses the log-derivative trick. The Gaussian *p*_0_ has no learned parameters and drops out, which gives Equation (2).

## B Learning Algorithm

### Algorithm 1

MLE learning of GLPM. The local state *ϕ*_i_ is the chain state 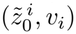 for BAOAB, with 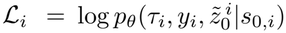 , or (*µ*_i_, *σ*_i_) for SVI, with *L*_i_ the ELBO of Equation (6).

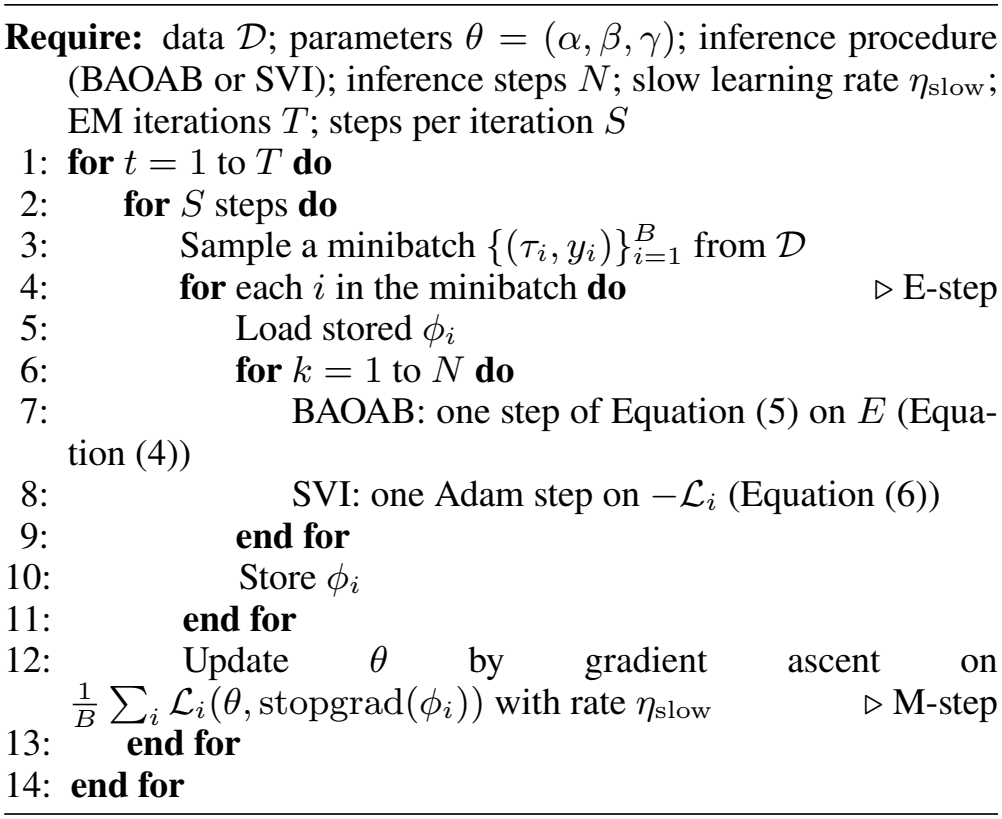

## C ICU-Sepsis Action Space

In ICU-Sepsis (Choudhary, Gupta, and Thomas 2024), each action specifies the dosages of intravenous (IV) fluids and vasopressors, discretized with thresholds similar to those of Komorowski et al. (2018). Table 6 lists the five dosage levels of each intervention; their combinations give the 25 discrete actions.

**Table 6:** Action space discretization. IV fluids and vasopressors are each binned into five dosage levels, yielding 5 × 5 = 25 treatment combinations. Level 0 denotes no administration.

| Level | IV fluids (mL/hr) | Vasopressor (μg/kg/min) |
|---|---|---|
| 0 | 0 | 0 |
| 1 | 0–49 | 0–0.08 |
| 2 | 50–149 | 0.08–0.20 |
| 3 | 150–499 | 0.20–0.45 |
| 4 | > 499 | > 0.45 |

## D Architecture and Training Details

Table 7 lists the hyperparameters of our model and Table 8 the planning configurations for the baselines. All offline methods share the same optimizer, schedule, and sequence-model backbone size; algorithm-specific hyperparameters use their official-implementation defaults.

**Table 7:** GLPM architecture and training settings. The five-step training and 25-step planning entries give the Langevin budget.

| Setting | Value |
|---|---|
| Latent dimension | 128 |
| Prior transformation | 1-D U-Net, hidden width 128 |
| Trajectory generator | 2-layer causal Transformer, 4 heads |
| Embedding width | 128 |
| Policy context | *K* = 10 steps |
| Survival predictor | 3-layer MLP, Bernoulli NLL |
| Shared optimizer | Adam |
| Shared learning rate | 10<sup>−4</sup> |
| Weight decay | 10<sup>−4</sup> |
| Batch size | 64 |
| Warmup | 10,000 steps |
| Total training | 500,000 steps (10 × 50,000) |
| Inference steps *N*, training | 5 |
| Inference steps *N*, planning | 25 |
| SVI learning rate | 0.3 |
| Hardware | One NVIDIA A6000 |

**Table 8:** Planning configurations for the baselines. Hyperpa-rameters not listed use the official-implementation defaults.

| Method | Setting | Value |
|---|---|---|
| CQL | Actor / critic learning rate | 0.025 |
| TT | Beam width / planning horizon | 256 / 15 |
| TT | Discretization bins / *k*<sub>act</sub> | 100 / 20 |
| Diffuser | Guidance scale / plan batch | 0 / 64 |
| DD | Classifier-free guidance scale | 1.5 |
| DD | Horizon / conditioning dropout | 8 / 0.1 |

## E Baseline Methods

### Behavior Cloning (BC)

BC trains a policy to mimic the actions of the behavior policy by minimizing the negative log-likelihood −E_*D*_[log *π*_ϑ_(*a*_t_|*s*_t*−*1_)] (Atkeson and Schaal 1997). It is computationally efficient but suffers from compounding errors when the learned policy deviates from the training distribution (Ross, Gordon, and Bagnell 2011).

### Conservative Q-Learning (CQL)

CQL mitigates overestimation in Q-function learning by penalizing Q-values for actions not well represented in the offline dataset (Kumar et al. 2020). Its conservative nature can lead to suboptimal policies when the dataset lacks high-quality actions (Uehara and Sun 2022).

### Decision Transformer (DT)

DT reframes RL as sequence modeling, predicting actions with a causal Transformer conditioned on past states, actions, and return-to-go (Chen et al. 2021). Return conditioning is unreliable when outcomes are stochastic and the return signal is weakly informative (Paster, McIlraith, and Ba 2022; Brandfonbrener et al. 2022); with binary, delayed rewards, the return-to-go carries little information for most of the trajectory.

### Q-Learning Decision Transformer (QDT)

QDT replaces DT’s return-to-go with Q-value estimates obtained by multi-step temporal-difference learning to improve credit assignment (Yamagata, Khalil, and Santos-Rodriguez 2023). Its reliance on Q-value estimation can introduce biases in sparse-reward settings.

### Trajectory Transformer (TT)

TT tokenizes states, actions, rewards, and returns-to-go by per-dimension quantile discretization, models them with a GPT-style Transformer, and plans by beam search scored by predicted return (Janner, Li, and Levine 2021). It inherits the noisy-signal problem of return-guided planning, and beam search adds substantial inference cost.

### Diffuser

Diffuser models entire state–action trajectories with a denoising diffusion model and steers sampling toward high return via value-function guidance (Janner et al. 2022), executing the first action of each sampled plan. Its iterative denoising makes planning computationally expensive.

### Decision Diffuser (DD)

DD replaces value guidance with classifier-free guidance on a target return (Ajay et al. 2023). On high-quality datasets with a narrow return distribution (e.g., Expert), classifier-free guidance is prone to cluster collapse: weak guidance leaves the sampler in a high-variance unconditional regime, while strong guidance amplifies the collapse, degrading performance (Table 1).

### Latent Plan Transformer (LPT)

LPT couples a latent prior, a causal Transformer trajectory decoder, and an outcome predictor under the factorization *p*(*z*) *p*(*τ* |*z*) *p*(*y*|*z*), inferring the latent plan at test time by MCMC from a target-conditioned posterior (Kong et al. 2024b). It is the most closely related baseline: it shares our use of a latent abstraction in place of return conditioning and differs from our model in the factorization.

## F EM Convergence across E-step Depths

Figure 7 shows the training negative log-likelihood and the posterior displacement over the ten EM iterations for each training E-step depth, as summarized in Section 4.4.

## G Posterior-Inference Chain Mixing

The EM diagnostics concern the training loop as a whole; they do not show whether a single BAOAB chain reaches its target within the inference budget. We run eight chains for 100 steps on the Medium-Replay checkpoint trained with *N* = 5, on the planning energy *E*_plan_ (Equation (8)) with *y*^⋆^ = 0.95 and *y*^⋆^ = 1.00. Each chain starts from its own draw of the Gaussian prior, so agreement between chains cannot come from a shared start. The energy needs no burnin for *y*^⋆^ = 0.95 and 13 steps for *y*^⋆^ = 1.00, both within the 25-step inference budget (Figure 8a). The per-step displacement ∥Δ*z*∥ stays near 1.55 throughout, so the chains keep moving (b). Chains from different starts agree: 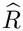 is 1.03 and 1.05 (d). Successive steps are strongly correlated: the energy autocorrelation reaches zero only after about 15 lags, and the effective-sample-size ratio is about 0.06 (c). Since planning keeps a single draw per episode, this correlation does not affect it.

**Figure 8:**
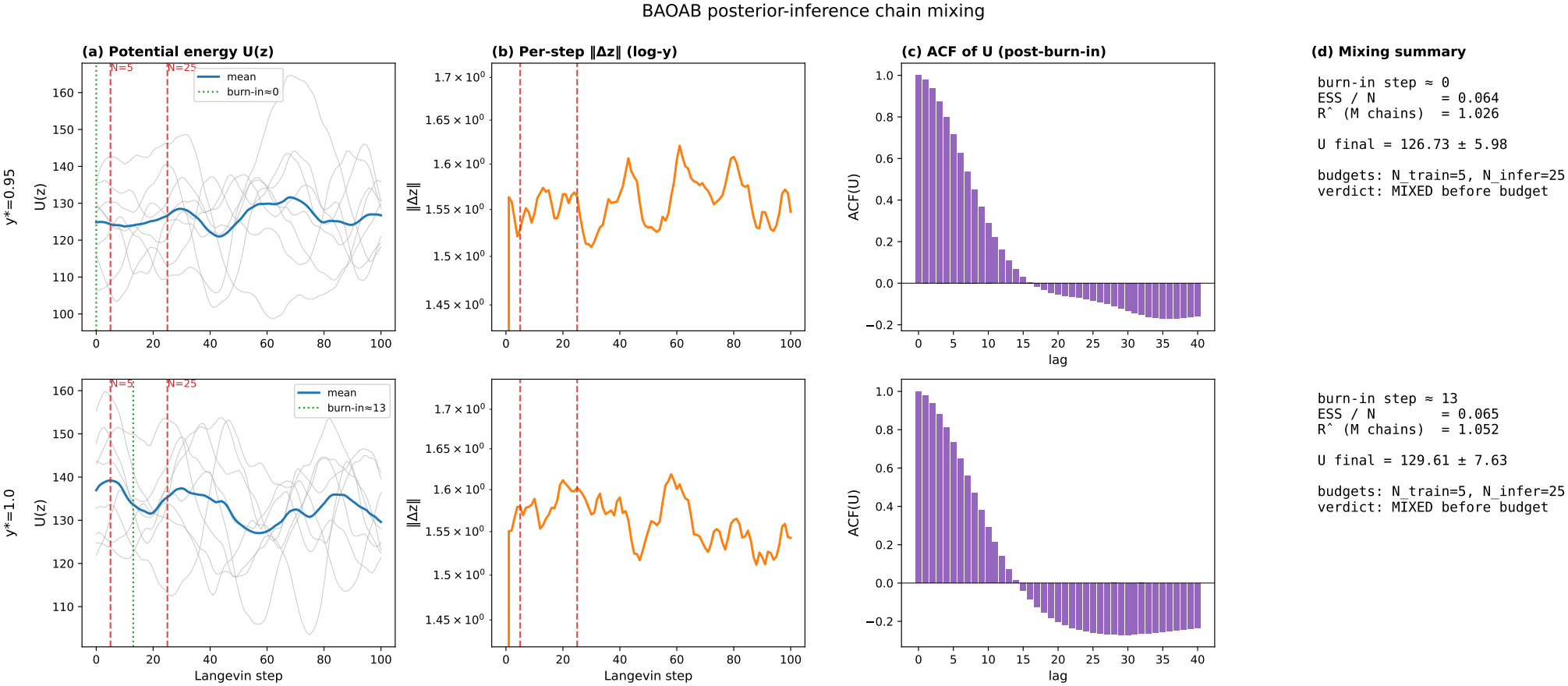
Mixing of eight BAOAB planning chains from diffuse prior samples on Medium-Replay, for *y*^⋆^ = 0.95 (top) and *y*^⋆^ = 1.00 (bottom). (a) Energy per chain; (b) per-step displacement ∥Δ*z*∥; (c) energy autocorrelation; (d) burn-in, ESS*/N* , and 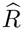.

## H Counterfactual Analysis Details

### H.1 Patient Selection and Methodology

For counterfactual analysis, we selected non-survivor patients from the test set with trajectory lengths ≥ 20 time steps to ensure sufficient temporal resolution for detecting treatment patterns. Patient 27 was selected as a representative case study for the main paper (trajectory length: 33 steps).

The counterfactual protocol was inferred using *N* = 25 Langevin dynamics steps with step size *h* = 0.1. Given a patient’s observed state sequence {*s*_0_, *s*_1_, … , *s*_H_}, we:

1. Infer a high-survival protocol *z*^⋆^ ∼ *p*_θ_(*z*_0_|*y*^⋆^ = 0.95) via Equation (8)
2. Generate counterfactual actions 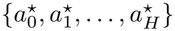 autoregressively using 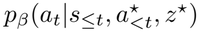
3. Compare with the clinician’s actual actions {*a*_0_, *a*_1_, … , *a*_H_}

We define *divergence* at time *t* as 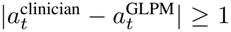 in *either* the vasopressor or fluid dimension. We additionally detect two clinically meaningful patterns using *dimension-specific* criteria:

- **De-escalation Prevention (vasopressors):** The clinician reduces vasopressor dosage at time *t* (i.e., 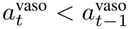), but GLPM maintains or increases support (i.e., 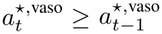).
- **Early Intervention (dimension-specific initiation):**
  - *Early vasopressor initiation:* 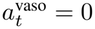 and 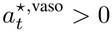.
  - *Early fluid initiation:* 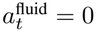 and 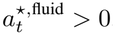.

### H.2 Time-Step Divergence Report (Patient 27)

Table 9 provides the complete intervention comparison for Patient 27, the representative case study shown in Figure 5.

**Table 9:**
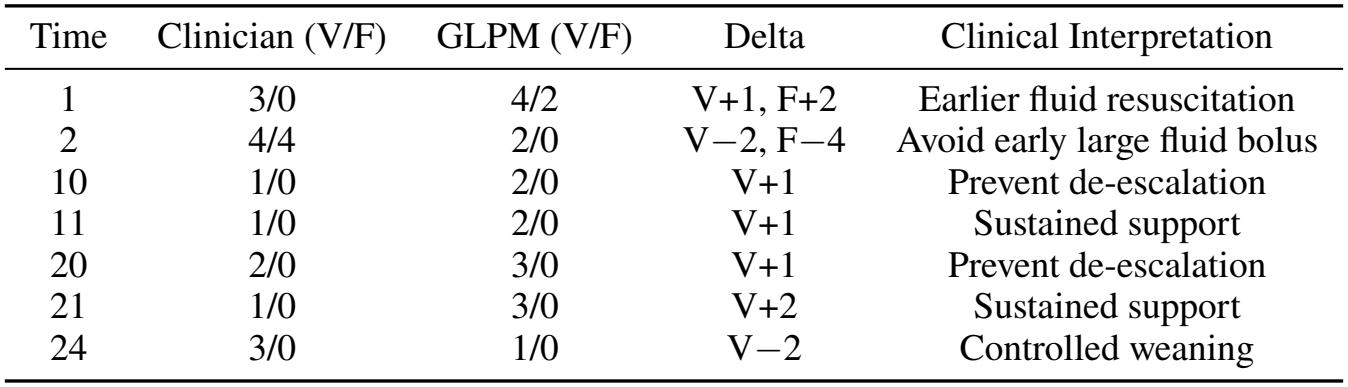
Time-step divergence report for Patient 27. V: Vasopressor level (0–4); F: Fluid level (0–4).

**Summary Statistics (Patient 27):**

- Total divergent steps: 7/33 (21.2%)
- Total vasopressor increases across divergent steps: +6 levels; net vasopressor change: +2 levels
- Net fluid change across divergent steps: −2 levels
- Mean absolute vasopressor divergence: 1.43 levels per divergent step

### H.3 Generalization Across Non-Survivors

To assess whether the patterns observed in Patient 27 generalize, we repeated the counterfactual analysis on *N* = 50 randomly sampled non-survivors (trajectory length ≥ 20 steps, random seed: 42). Table 10 summarizes the aggregate results. Figure 9 shows the distribution of each metric across the 50 analyzed patients.

**Table 10:** Counterfactual analysis across non-survivors (N=50).

| Metric | Mean ± Std | Interpretation |
|---|---|---|
| Divergence rate | 24.2% ± 9.8% | Consistent selective intervention |
| Net vasopressor change | +1.3± 4.5 levels | GLPM favors sustained support |
| Net fluid change | +0.1± 2.9 levels | Neutral fluid strategy |
| De-escalation preventions | 1.8 ± 1.5 per patient | Primary divergence mechanism |

**Figure 9:**
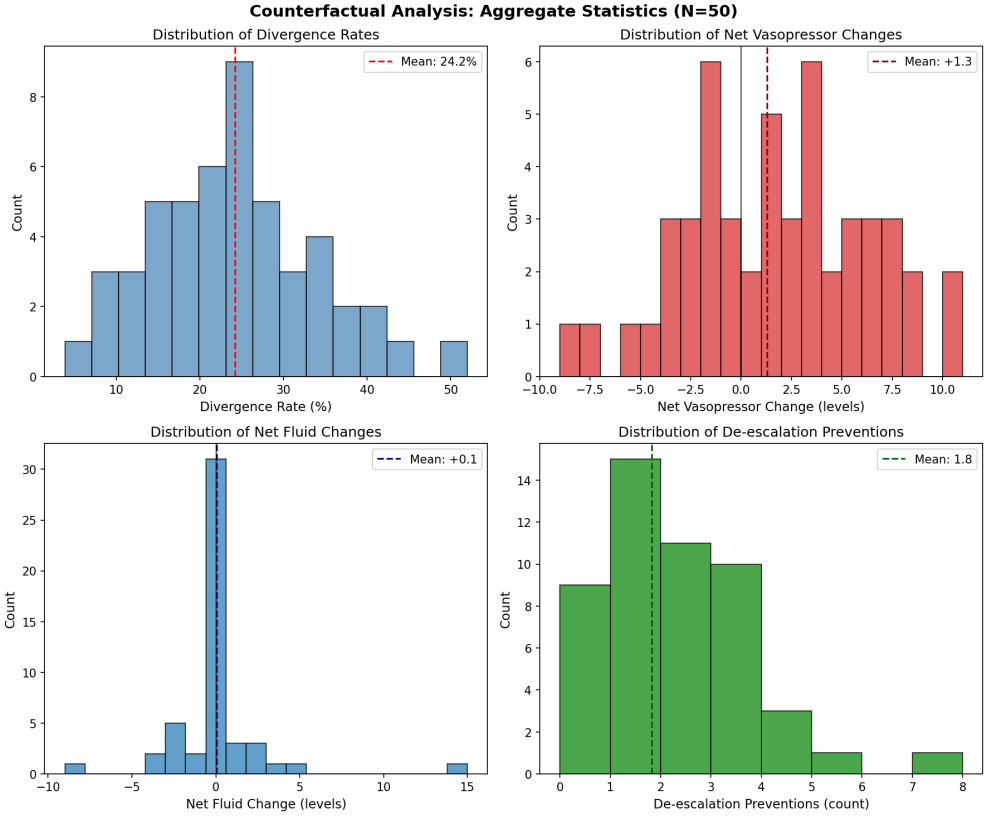
Distribution of counterfactual analysis metrics across *N* = 50 non-survivors. **(Top Left)** Divergence rates are concentrated around 20–30%, indicating GLPM makes selective rather than wholesale changes. **(Top Right)** Net vasopressor changes are right-skewed with mean +1.3 levels, confirming GLPM’s preference for sustained hemodynamic support. **(Bottom Left)** Net fluid changes are centered around zero, suggesting fluid management is patient-specific rather than systematic. **(Bottom Right)** De-escalation prevention events occur ≈1–2 times per patient on average, representing the primary mechanism of divergence.

**Figure 10:**
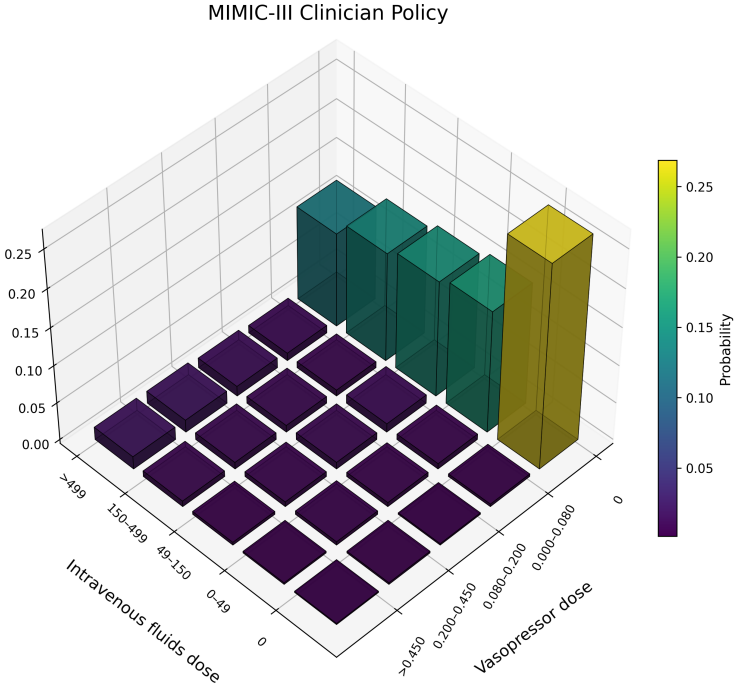
Action distribution of the clinician policy from MIMIC-III. Counts are aggregated across all timesteps over the discretized dose bins for IV fluids and vasopressors, where level 0 denotes no administration.

**Figure 11:**
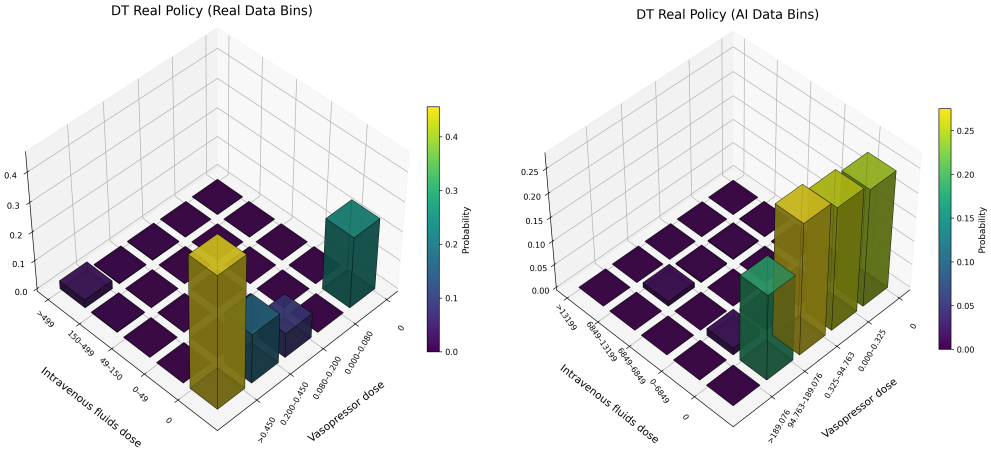
Action distribution of the DT policy trained on real MIMIC trajectories. Left: bins defined by clinician-derived dose thresholds. Right: bins defined by DT-derived dose thresholds. Counts are aggregated across all timesteps; level 0 denotes no administration.

**Figure 12:**
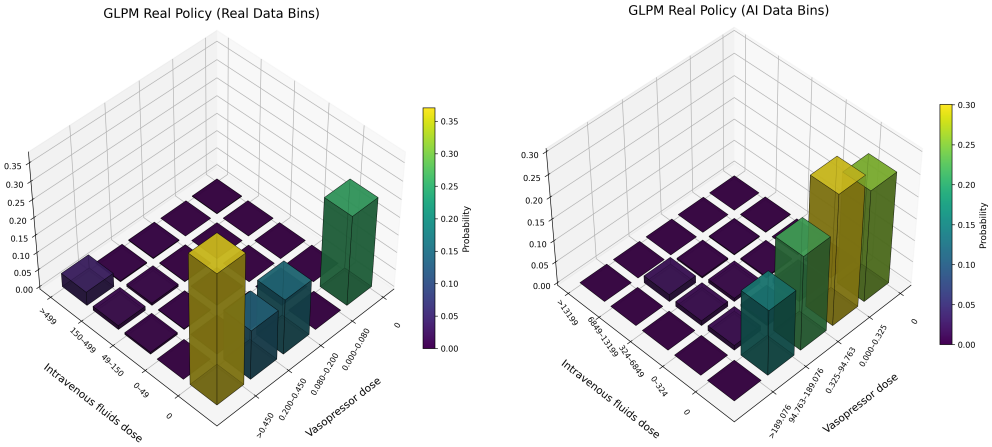
Action distribution of the GLPM policy trained on real MIMIC trajectories. Left: bins defined by clinician-derived dose thresholds. Right: bins defined by GLPM-derived dose thresholds. Counts are aggregated across all timesteps; level 0 denotes no administration.

**Figure 13:**
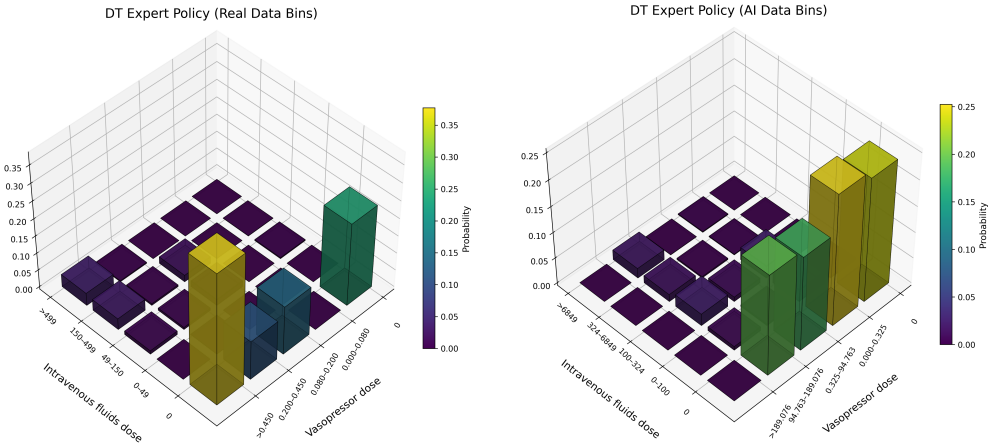
Action distribution of the DT policy trained on the *Expert* offline dataset. Counts are aggregated across all timesteps over five discretized dose bins for IV fluids and vasopressors (level 0 denotes no administration). Left: bins defined using clinician-derived dose thresholds. Right: bins defined using DT-derived dose thresholds.

**Figure 14:**
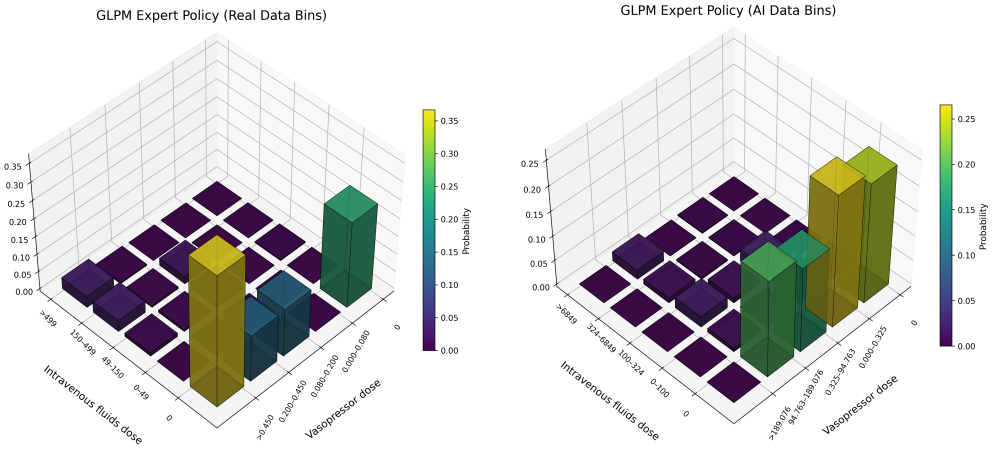
Action distribution of the GLPM policy trained on the *Expert* offline dataset. Counts are aggregated across all timesteps over five discretized dose bins for IV fluids and vasopressors (level 0 denotes no administration). Left: bins defined using clinician-derived dose thresholds. Right: bins defined using GLPM-derived dose thresholds.

#### Key Findings

Across *N* = 50 non-survivors, GLPM makes selective edits rather than wholesale policy replacement (divergence rate: 24.2% ± 9.8%). On average, it recommends higher vasopressor support (net change: +1.3 ± 4.5 levels), while fluid recommendations remain near-neutral (net change: +0.1 ± 2.9 levels), indicating that increased hemodynamic support is not paired with uniformly more aggressive fluids. De-escalation prevention occurs 1.8 ± 1.5 times per patient on average, consistent with temporal smoothing of vasopressor support. These aggregate results align with the Patient 27 case study.

### H.4 Limitations

We note several limitations of this counterfactual analysis:

- **Simulation Environment:** All counterfactual trajectories are generated within the ICU-Sepsis simulation. The recommended actions have not been validated in real clinical settings or with actual patient physiology.
- **Fixed State Sequences:** We replay the patient’s *observed* state sequence under the counterfactual protocol. In reality, different actions would lead to different subsequent states, so these results represent “what GLPM would have recommended” rather than “what would have happened.”
- **Selection Bias:** We analyze only non-survivors with sufficiently long trajectories. The patterns identified may not generalize to shorter stays or survivors.
- **Causal Interpretation:** While we use the term “counterfactual,” these analyses do not establish causal efficacy. Prospective validation would be required to determine whether GLPM’s recommendations actually improve outcomes.

## I Inference Runtime

We benchmarked BAOAB inference (*N* = 25 Langevin steps) over 100 episodes on a single GPU (Table 11). Protocol inference is a one-time cost per episode; subsequent action generation is autoregressive. A full episode averages 1.18 s. Since ICU-Sepsis decisions are made in 4-hour blocks, this is about 0.008% of the decision interval. Langevin planning adds latency relative to single-forward-pass baselines such as BC or DT, but the difference is negligible at the clinical decision cadence.

**Table 11:**
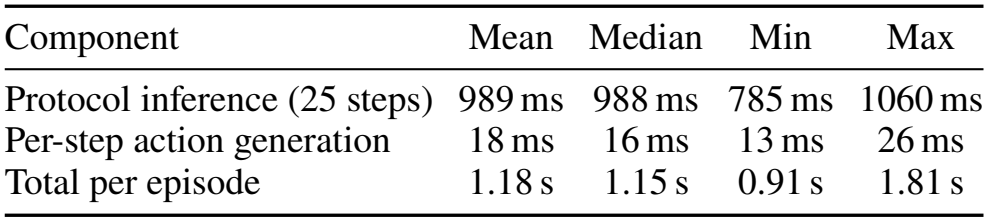
BAOAB inference wall-clock over 100 episodes on one GPU.

## J Off-Policy Evaluation on Real Trajectories

Survival on Real is measured by simulator rollouts of policies trained on MIMIC-III trajectories mapped into the ICU-Sepsis MDP (Choudhary, Gupta, and Thomas 2024), consistent with all baselines. To complement it, we computed self-normalized weighted importance sampling (WIS) on held-out Real trajectories with bootstrap 95% confidence intervals (10,000 resamples; Table 12). WIS recovers the empirical clinician survival rate (0.755) exactly, since all importance ratios equal one when the evaluation and behavior policies coincide; this validates the implementation. For learned policies, cumulative importance ratios collapse over multi-step trajectories, producing intervals that span nearly [0, 1]. This is a well-documented limitation of importance sampling in sepsis treatment (Raghu et al. 2017; Komorowski et al. 2018; Jeter et al. 2019): learned policies recommend systematically different treatments from clinicians (Figure 6), so few trajectories carry non-negligible weight. Simulator-based evaluation remains the standard on this benchmark for this reason.

**Table 12:** Simulator survival and WIS estimates on held-out Real trajectories, with bootstrap 95% CIs.

| Policy | Simulator | WIS | WIS 95% CI |
|---|---|---|---|
| Clinician | 0.755 | 0.755 | [0.742, 0.768] |
| CQL | 0.784 | 1.000 | [0.738, 1.000] |
| DT | 0.802 | 0.194 | [0.000, 0.964] |
| LPT | 0.802 | 0.379 | [0.000, 1.000] |
| GLPM (SVI) | 0.820 | 0.874 | [0.000, 1.000] |
| GLPM (BAOAB) | 0.832 | 0.127 | [0.000, 0.999] |

## K Trained Policy Action Distributions

We visualized the action distributions of DT and GLPM, as well as the actual clinician policy.

## Footnotes

1 At *K* = 30, three times the mean length, BAOAB reaches 86.0 *±*3.8% and DT 81.2 *±*2.8%. We leave this setting out because most of its window is padding.

